# Efficacy of Botulinum Toxin Type A in Reducing Facial Wrinkles: A Comprehensive Review of Clinical Outcomes

**DOI:** 10.1101/2025.05.14.25327581

**Authors:** Reza Ghalamghash

**Affiliations:** Premium College, Toronto, Canada

**Keywords:** Botulinum Toxin A, Facial Wrinkles, Cosmetic Dermatology, Clinical Efficacy, Literature Review

## Abstract

**Objective:** This literature review synthesizes current evidence regarding the efficacy and safety of Botulinum Toxin Type A (BoNT-A) for the treatment of facial wrinkles. The increasing demand for minimally invasive cosmetic procedures has positioned BoNT-A as a leading intervention for managing facial rhytids.

**Methods:** This review examines clinical outcomes across various facial areas, including glabellar lines, crow’s feet, and forehead lines, considering different BoNT-A formulations, dosages, and injection techniques. The methodology involved a comprehensive search of major electronic databases for studies published between 2014 and 2024.

**Results:** Findings indicate that BoNT-A is consistently effective in reducing the severity of dynamic facial wrinkles, with high patient satisfaction reported across different treatment areas and formulations. While generally safe, potential adverse events such as eyelid ptosis and the risk of immunogenicity with repeated use are important considerations.

**Conclusion:** The review highlights the need for ongoing research to optimize treatment protocols, explore long-term effects, and compare the efficacy of different BoNT-A products and emerging alternatives.

## 1. Introduction

### 1.1 Background on Facial Wrinkles and the Growing Demand for Non-Surgical Cosmetic Treatments

Facial wrinkles are a visible manifestation of skin aging, often associated with repetitive muscle contractions during facial expressions (1). These lines can impact an individual’s self-perception and how others perceive them, sometimes leading to feelings of being misinterpreted as angry, anxious, or fatigued. In response to the desire for a more youthful appearance, there has been a significant increase in the demand for aesthetic interventions. Among these, minimally invasive procedures have gained considerable popularity due to their reduced downtime and lower risk profiles compared to surgical options (1, 2). Notably, a significant portion of individuals seeking these treatments are women between the ages of 35 and 50.4 This growing acceptance and availability of aesthetic treatments underscore the importance of understanding the efficacy and safety of various interventions, particularly Botulinum Toxin Type A (BoNT-A), which has become a cornerstone in non-surgical cosmetic enhancement (1).

### 1.2 Overview of Botulinum Toxin Type A and Its Mechanism of Action in Reducing Wrinkles

Botulinum toxin is a neurotoxic protein derived from the bacterium Clostridium botulinum. Seven distinct serotypes of botulinum toxin exist (A-G), with type A being the most commonly utilized in aesthetic medicine. The mechanism by which BoNT-A reduces facial wrinkles involves the temporary paralysis of underlying muscle activity. This is achieved by inhibiting the release of acetylcholine, a neurotransmitter responsible for muscle contraction, at the neuromuscular junction. The toxin binds to the nerve terminal, is internalized, and then interferes with the proteins necessary for the fusion of acetylcholine-containing vesicles with the nerve cell membrane. This process ultimately prevents muscle contraction, leading to a smoothing of dynamic wrinkles, which are those that appear during muscle movement (3, 4). The effects of BoNT-A typically become noticeable within 24 to 72 hours after injection, reaching maximum effect around one to four weeks, and generally last for approximately 8 to 12 weeks. Several commercially available formulations of BoNT-A exist, including onabotulinumtoxinA, abobotulinumtoxinA, incobotulinumtoxinA, daxibotulinumtoxinA, prabotulinumtoxinA, letibotulinumtoxinA, and relabotulinumtoxinA (4, 5). While all these formulations contain botulinum toxin type A, they may differ in their molecular weight, potency units, and manufacturing processes, making them non-interchangeable (4).

### 1.3 Significance of BoNT-A in Aesthetic Medicine and the Need for a Comprehensive Review

Botulinum Toxin Type A has become one of the most popular and widely used non-surgical cosmetic procedures for the management of facial rhytids. Its effectiveness and relatively favorable safety profile have contributed to its widespread adoption in aesthetic medicine. As a minimally invasive procedure with minimal downtime, BoNT-A injections offer a convenient option for individuals seeking to address facial lines caused by muscle hyperactivity (6). The field of aesthetic medicine is continuously evolving, with ongoing advancements in BoNT-A formulations and injection techniques (7). New botulinum toxin products, often termed “ biosimilars,” along with innovative formulations of established BoNT types, are continuously emerging. Given the widespread use and ongoing developments in this area, it is crucial to have a comprehensive and up-to-date understanding of the clinical evidence supporting the efficacy and safety of BoNT-A for facial wrinkle reduction (8). In the increasing accessibility of aesthetic treatments and the growing volume of information available to patients, platforms such as https://premiumdoctors.org/ may serve as resources to connect individuals with healthcare professionals specializing in aesthetic procedures. These platforms aim to provide a convenient avenue for patients to find qualified medical practitioners who can offer guidance and administer treatments like BoNT-A injections. Resources like https://premiumdoctors.org/ strive to connect individuals with such professionals who possess the necessary expertise to provide safe and effective cosmetic interventions.

This review aims to synthesize the current scientific literature to provide clinicians and researchers with an evidence-based overview of the clinical outcomes associated with BoNT-A treatment for facial wrinkles.

### 1.4 Objectives of the Current Literature Review

The primary objectives of this literature review are to comprehensively evaluate the efficacy of Botulinum Toxin Type A in reducing facial wrinkles, specifically focusing on glabellar lines, crow’s feet, and forehead lines. This review also aims to assess the safety profile of BoNT-A in these cosmetic applications by examining the incidence and nature of reported adverse events. Furthermore, it seeks to identify existing gaps in the current knowledge and suggest potential directions for future research in this continually evolving field of aesthetic medicine.

## 2. Methodology

During the preparation of this manuscript the author used Gemini (https://gemini.google.com/) in order to collect information and writing articles. After using this tool/service, the author physically reviewed and edited the content as needed and take full responsibility for the content of the publication.

### 2.1 Search Strategy

A comprehensive search of relevant scientific literature was conducted using the following electronic databases: PubMed/MEDLINE (https://www.nlm.nih.gov/medline/), Scopus (https://www.scopus.com/), Web of Science (https://www.webofscience.com/), and the Cochrane Library (https://www.cochrane.org/). The search strategy employed a combination of key terms and Boolean operators to identify studies relevant to the efficacy of Botulinum Toxin Type A in treating facial wrinkles. The primary search terms included “ Botulinum Toxin Type A,” “ facial wrinkles,” “ cosmetic use,” “ aesthetic,” “ clinical outcomes,” “ efficacy,” “ safety,” “ systematic review,” “ meta-analysis,” “ glabellar lines,” “ frown lines,” “ crow’s feet,” “ lateral canthal lines,” “ forehead lines,” and “ horizontal forehead lines” (9, 10). The search was limited to studies published in the English language between January 2014 and December 2024 to ensure the inclusion of recent and relevant research.

### 2.2 Article Selection Process

The identified articles underwent a rigorous selection process based on predefined inclusion and exclusion criteria. Inclusion criteria comprised studies published in peer-reviewed journals between 2014 and 2024 that focused on the aesthetic use of BoNT-A for treating facial wrinkles. This included clinical trials (randomized controlled trials, non-randomized controlled trials), systematic reviews, and meta-analyses that reported on clinical outcomes, efficacy (e.g., responder rates, improvement scales), and safety (e.g., adverse events). Studies involving adult human participants were also included. Exclusion criteria consisted of studies published before 2014, studies focusing solely on therapeutic (non-cosmetic) applications of BoNT-A (unless relevant for context or mechanism), animal studies, editorials, letters to the editor, non-peer-reviewed publications, studies not providing sufficient data on efficacy or safety, and studies not published in English. The article screening process involved reviewing titles and abstracts to assess initial eligibility, followed by a full-text review of potentially relevant articles to confirm adherence to the inclusion and exclusion criteria. Data extraction was performed to gather information on study design, participant demographics, BoNT-A formulation and dosage, injection techniques, outcome measures, key findings related to efficacy and safety, and reported adverse events. To ensure the reliability of the selection process, it is recommended that two independent reviewers conduct screening and data extraction. The number of articles initially identified, screened, and finally included in this review will be detailed in the full publication.

## 3. Findings

### 3.1 Efficacy of BoNT-A for Glabellar Lines

The efficacy of BoNT-A for treating glabellar lines, also known as frown lines, has been extensively studied in numerous clinical trials and systematic reviews (11, 12). Meta-analyses have consistently demonstrated significantly higher effective rates in BoNT-A treatment groups compared to placebo (13). Various formulations, including onabotulinumtoxinA, abobotulinumtoxinA, incobotulinumtoxinA, daxibotulinumtoxinA, letibotulinumtoxinA, and relabotulinumtoxinA, have shown significant improvement in the appearance of glabellar wrinkles (11). Responder rates, defined as a significant improvement in wrinkle severity, are typically high across different studies. For instance, a phase 3 clinical trial evaluating a new BoNT-A formulation, BMI2006, reported response rates of 78.83% in the BMI2006 group and 83.09% in the onabotulinumtoxinA group at week 4, demonstrating non-inferiority between the two (14). The standard dosage for treating glabellar lines is generally 20 units. Research suggests that increasing the dose beyond the standard might prolong the duration of the treatment effect without a significant increase in safety concerns. Median durations of around 6 to 9 months have been reported with higher doses of different BoNT-A formulations (14, 15). Comparative studies between different BoNT-A products for glabellar lines often indicate comparable efficacy. For example, a study comparing incobotulinum toxin A and abobotulinum toxin A found no statistically significant difference in improvement rates at week 4, although abobotulinum toxin A showed a higher improvement rate at week 24 (15). Similarly, BMI2006 was found to be as effective as onabotulinumtoxinA in improving glabellar wrinkles in Asian adults (14).

**Table 1:** Summary of Key Studies on BoNT-A Efficacy for Glabellar Lines.

| **Study**<br>**(Citation)** | **BoNT-A**<br>**Formulation**<br>**and**<br>**Dosage** | **Number of**<br>**Participants** | **Key**<br>**Findings**<br>**(Responder**<br>**Rate at**<br>**Week 4)** | **Duration of**<br>**Effect** | **Snippet**<br>**ID(s)** |
| --- | --- | --- | --- | --- | --- |
| Camargo et al. (2021) | Various<br>(Meta-analysis) | 14,919 | Higher success rates vs. placebo | ~20 weeks | (16) |
| Guo et al. (2015) | OnabotulinumtoxinA 20 U | 1474 | Significantly higher efficacy vs. placebo | Not specified | (17) |
| Choi et al. (2024) | BMI2006 20 U;<br>OBoNT 20 U | 276 | 78.83% (BMI2006),<br>83.09% (OBoNT) | 8-12 weeks | (14) |
| Han et al. (2021) | MBA-P01 20 U; ONA-BoNT-A 20 U | 318 | 87.42% (MBA-P01),<br>82.91% (ONA-BoNT-A) | Decreases over time | (18) |
| Fabi et al. (2021) | ABO (High Dose); INCO (High Dose); ONA (High Dose) | Various | Prolonged duration with higher doses | Up to 9 months | (19) |
| Chan et al. (2019) | IncobotulinumtoxinA 12-20 U | 45 | 100% response rate at Day 14 | At least 4 months | (20) |
| Kestemont et al. (2022) | AboBoNT-A 50 U; INCO 20 U; ONA 20 U | Not specified | High responder rates | Not specified | (21) |
| Sforzolini et al. (2022) | RelabotulinumtoxinA | >1900 | Significant improvement vs. placebo | ~6 months | (22) |
| Gold et al. (2024) | LetibotulinumtoxinA | Not specified | Higher response in females 35-50 | Up to Week 16 | (23) |

### 3.2 Efficacy of BoNT-A for Crow’s Feet

The use of BoNT-A for the treatment of lateral canthal lines, commonly known as crow’s feet, has also demonstrated significant efficacy (24). Clinical trials have shown that onabotulinumtoxinA results in significantly greater responder rates compared to placebo in reducing the severity of crow’s feet lines (24). Similarly, relabotulinumtoxinA has been found to significantly improve the appearance of crow’s feet, whether administered alone or in conjunction with treatment for glabellar lines (25). The typical dosage for onabotulinumtoxinA in treating crow’s feet is around 24 units, often administered using specific injection patterns (26). Injection landmarks are commonly recommended at a point 1.5 to 2.0 cm lateral to the lateral canthus. Studies comparing different injection techniques, such as a 3-point versus a 4-point intramuscular injection, suggest that adding a fourth injection point may offer additional benefits, particularly for patients with lower-fan crow’s feet patterns, and can lead to higher patient satisfaction without a significant increase in adverse events. The duration of the treatment effect for crow’s feet typically ranges from 3 to 4 months (27). Clinical studies have reported median response durations for onabotulinumtoxinA in the range of 119 to 148 days (28).

**Table 2:** Summary of Key Studies on BoNT-A Efficacy for Crow’s Feet.

| **Study**<br>**(Citation**<br>**)** | **BoNT-A**<br>**Formulat**<br>**ion and**<br>**Dosage** | **Number**<br>**of**<br>**Participa**<br>**nts** | **Key**<br>**Findings**<br>**(Respond**<br>**er Rate**<br>**at Week**<br>**4)** | **Duration**<br>**of Effect** | **Injection**<br>**Techniqu**<br>**e** | **Snippet**<br>**ID(s)** |
| --- | --- | --- | --- | --- | --- | --- |
| Camargo<br>et al.<br>(2021) | Various<br>(Meta-analysis) | 14,919 | Effective<br>in<br>reducing<br>crow's<br>feet | ~20<br>weeks | Not<br>specified | (16) |
| Baumann et al. (2016) | Onabotulinumtoxin A 24 U | 1362 | ≥1-grade improvement in FWS scores | ~4 months | Not specified | (28) |
| Carruthers et al. (2016) | Onabotulinumtoxin A 12 U, 24 U | 1362 | Significantly greater response vs. placebo | Not specified | 2 injection patterns | (29) |
| Quanget al. (2024) | BoNTA (various units) | 60 | Significant decrease in CFGS score | ~16 weeks | 3-point vs. 4-point | (27) |
| Galderma Press Release (2022) | Relabotulinumtoxin A | >1900 | Significant improvement vs. placebo | ~6 months | Not specified | (22) |

### 3.3 Efficacy of BoNT-A for Forehead Lines

The treatment of horizontal forehead lines with BoNT-A has also been shown to be effective in reducing their appearance (7). However, this treatment area presents a challenge due to the risk of inducing brow ptosis (drooping of the eyebrows) if the frontalis muscle, responsible for elevating the brows, is inadvertently affected. Traditional injection techniques for forehead lines typically recommend placing injection points approximately 1.5 to 2 cm above the orbital rim to minimize this risk. To optimize efficacy and minimize the risk of brow ptosis, various refined and novel injection techniques have been developed and studied. One such technique is the Lines and Dots (LADs) technique, which aims to improve the precision of BoNT-A administration by targeting specific anatomical landmarks related to the frontalis muscle and the supraorbital and supratrochlear nerves. This technique has been associated with high patient satisfaction rates and a low incidence of side effects (30). Another approach, known as Microbotox, involves using multiple small injections of diluted BoNT-A across the lower frontalis muscle. This technique has shown promise in treating mild forehead wrinkles and rhytides without causing significant changes in brow position (30). Studies have also explored the use of hyperdiluted incobotulinumtoxinA for treating forehead lines, reporting significant improvement in line severity with sustained results over several months (31). The typical dosage for treating forehead lines can vary depending on the formulation and technique used, with some studies reporting effective results with around 10 units of hyperdiluted incobotulinumtoxinA (31). The duration of effect for forehead line treatment is generally similar to other facial areas, typically lasting for approximately 3 to 4 months, although some studies have reported sustained improvement for longer periods (32).

**Table 3:**
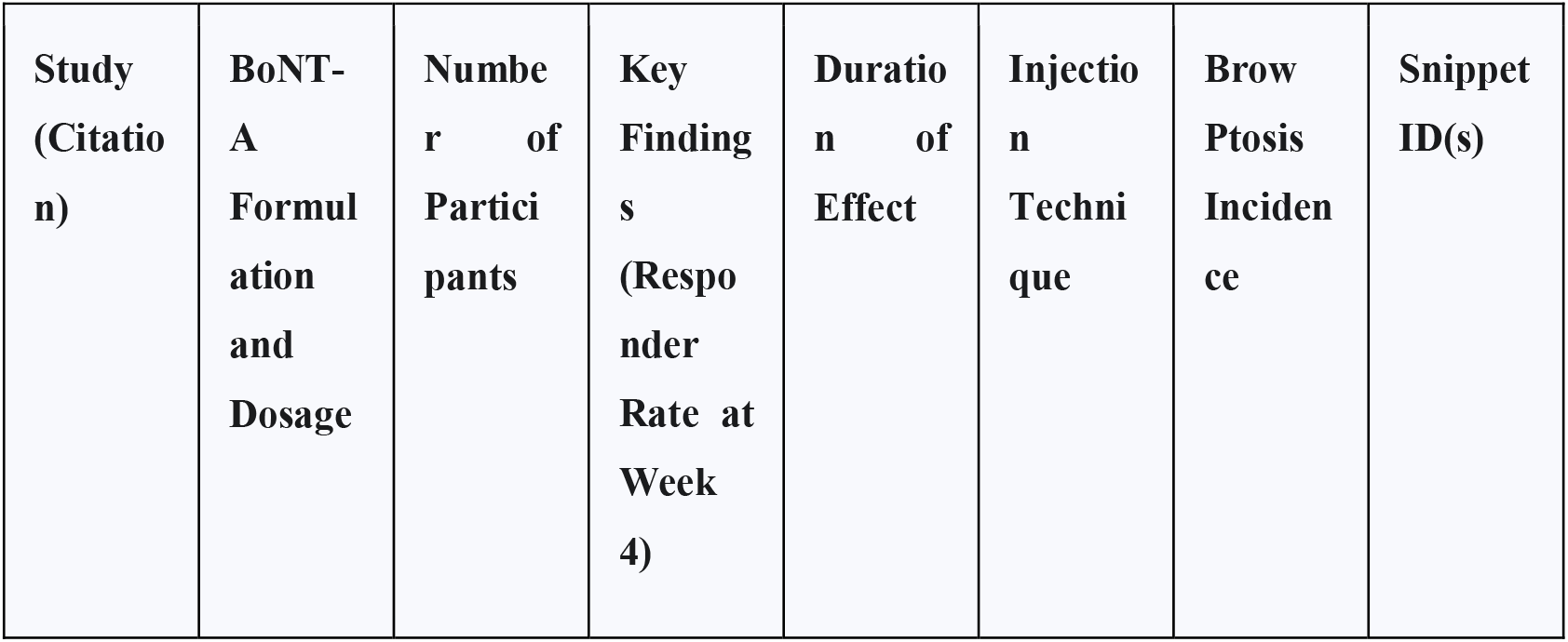
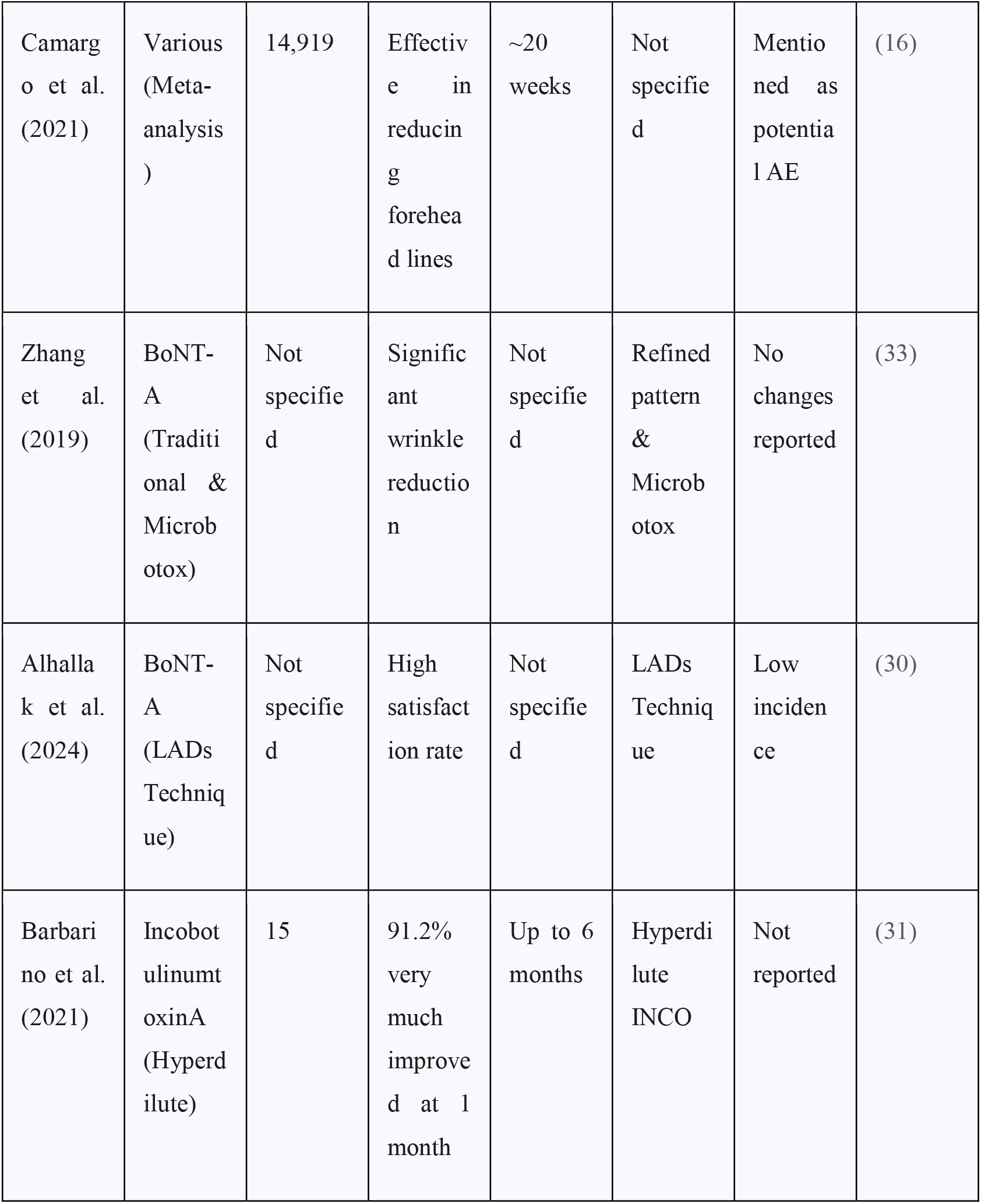
Summary of Key Studies on BoNT-A Efficacy for Forehead Lines.

### 3.4 Overall Duration of Effect

The duration of the clinical efficacy of BoNT-A in treating facial wrinkles generally ranges from approximately 3 to 6 months. A meta-analysis encompassing a large number of studies reported a mean duration of treatment effect of around 20 weeks (34). Several factors can influence how long the effects of BoNT-A last, including the dosage administered, the specific injection technique employed, an individual’s metabolism, and the initial severity of the wrinkles being treated (35). Research has also explored the possibility of extending the duration of BoNT-A’s effects by using higher doses than the standard recommendations (36). For instance, studies on glabellar lines have indicated that a 2 to 2.5-fold increase in the on-label dose of abobotulinumtoxinA or a 5-fold increase in the incobotulinumtoxinA dose can potentially prolong the duration of response to around 9 months (4). Similarly, a 2 to 4-fold increase in the onabotulinumtoxinA on-label dose has been reported to yield a median duration of approximately 6 months (4). These findings suggest that while the typical duration is within the 3 to 6-month window, adjusting the dosage for certain formulations might offer the possibility of less frequent treatments for some patients.

### 3.5 Safety and Adverse Events

Botulinum Toxin Type A is generally considered a safe treatment for facial wrinkles; however, like any medical procedure, it is associated with potential adverse events (37). The most common side effects are typically mild and transient, including pain, bruising, swelling, and erythema at the injection sites, as well as headache (38). More significant, though less frequent, adverse events can occur, such as eyelid ptosis (drooping), brow ptosis, and facial asymmetry (39). Meta-analyses suggest that the risk of major adverse events, particularly eyelid ptosis, is probably increased with BoNT-A treatment compared to placebo (40). In rare cases, the effects of the toxin can spread beyond the injection site, potentially leading to more serious complications such as swallowing and breathing difficulties (41). Another important consideration is the immunogenicity of BoNT-A. Repeated injections can potentially trigger the formation of neutralizing antibodies, which may reduce the effectiveness of the treatment over time (42). Therefore, while BoNT-A is generally safe for cosmetic use, it is crucial for practitioners to be aware of these potential adverse events and to take appropriate measures to minimize their occurrence.

### 3.6 Patient Satisfaction

Patient satisfaction is a critical outcome measure in aesthetic treatments, and studies have consistently reported high levels of satisfaction with BoNT-A injections for facial wrinkles across various treatment areas (43). High satisfaction is often attributed to the visible reduction in wrinkle severity and the resulting more youthful appearance (43). Studies utilizing patient-reported outcome measures have further supported these findings. For example, in a study on crow’s feet, subjective patient-rated satisfaction scores were significantly higher in the group receiving a 4-point injection technique compared to those receiving a 3-point injection (27). Similarly, studies on incobotulinumtoxinA have shown that patient satisfaction ratings remain high throughout the treatment effect (44). Achieving a natural-looking result is also a significant factor contributing to patient satisfaction, and techniques like using hyperdiluted incobotulinumtoxinA have been reported to help achieve this desired outcome. Overall, the consistently high levels of patient satisfaction indicate that BoNT-A is generally effective in meeting patient expectations for facial wrinkle reduction and aesthetic improvement.

## 4. Discussion

The findings from the reviewed literature consistently demonstrate the efficacy of BoNT-A in reducing facial wrinkles across the glabellar, crow’s feet, and forehead regions. The evidence base is robust, with numerous randomized controlled trials and systematic reviews supporting its use. While different BoNT-A formulations generally exhibit comparable effectiveness in reducing wrinkle severity, some variations exist in the likelihood of specific adverse events, such as eyelid ptosis, and the potential duration of the treatment effect. Optimal dosages and injection techniques vary depending on the facial area being treated and the specific BoNT-A formulation used. Refined injection techniques, particularly for forehead lines and crow’s feet, have been shown to improve efficacy and minimize the risk of complications like brow ptosis.

A significant strength of the available literature is the large number of randomized controlled trials and systematic reviews, providing a substantial body of evidence supporting the efficacy of BoNT-A for facial wrinkles. However, the reviewed literature also presents certain limitations. Heterogeneity in study designs, including variations in the scales used for wrinkle assessment and differences in follow-up periods, can make direct comparisons across studies challenging and can hinder comprehensive meta-analyses. Additionally, the potential for publication bias, where studies with positive results are more likely to be published, should be considered. There is also a need for more long-term studies to fully understand the effects of repeated BoNT-A treatments and the development of immunogenicity. Furthermore, research including more diverse ethnicities and genders is needed to ensure the generalizability of the findings across different populations.

Several gaps remain in our current understanding of BoNT-A for facial wrinkles. The long-term effects of repeated BoNT-A treatments on muscle structure and function are not fully elucidated. More data is needed on the efficacy and safety of newer BoNT-A formulations in diverse patient populations. There is also limited evidence on the optimal intervals for retreatment based on individual patient factors and the potential for prolonged effects with specific formulations or dosages. Finally, more research is needed to directly compare BoNT-A with other emerging aesthetic treatments, including non-invasive alternatives like peptides and topical formulations, to better understand their relative efficacy and long-term outcomes.

Future research should focus on addressing the identified gaps in knowledge. Randomized controlled trials directly comparing different BoNT-A products head-to-head, using standardized outcome measures and including long-term follow-up, are needed to assess sustained efficacy and safety. Further investigation into optimal dosages and injection techniques for diverse patient populations, considering factors such as age, sex, muscle mass, and specific wrinkle patterns, would be valuable. Studies examining the long-term safety and efficacy of repeated BoNT-A treatments, the incidence and clinical significance of immunogenicity, and strategies to mitigate antibody formation are also warranted. Finally, research comparing the efficacy, safety, cost-effectiveness, and patient satisfaction of BoNT-A with emerging non-invasive alternatives would provide valuable insights for clinicians and patients.

## 5. Conclusion

This comprehensive literature review confirms the established efficacy and acceptable safety profile of Botulinum Toxin Type A for the temporary reduction of facial wrinkles in the glabellar, crow’s feet, and forehead regions. The evidence supports the use of various BoNT-A formulations for achieving significant aesthetic improvements with high levels of patient satisfaction. However, clinicians should adopt individualized treatment approaches based on patient needs, specific wrinkle characteristics, and the properties of different BoNT-A formulations, while also carefully considering potential adverse events and the temporary nature of the results. Ongoing research is essential to address existing knowledge gaps, optimize treatment protocols, and further explore the long-term effects and comparative effectiveness of BoNT-A in the dynamic field of aesthetic medicine.

## Data Availability

All data produced in the present work are contained in the manuscript

## Acknowledgements

(if necessary)

